# Role of accuracy measures in selecting hepatocellular carcinoma patients for liver transplantation

**DOI:** 10.1101/2024.12.16.24319098

**Authors:** Laura P. Frazão, José B. Pereira-Leal, Christophe Duvoux, Joana Cardoso

## Abstract

**Importance:** Multiple criteria are used worldwide to select hepatocellular carcinoma (HCC) patients with a low risk of recurrence for liver transplantation (LT). However, it remains unclear which criteria are best for the LT-involved stakeholders, particularly in accurately identifying patients at high risk of recurrence.

**Objective:** To identify the most accurate criteria for selecting HCC patients for LT.

**Data Sources:** In June 2023, a systematic literature search was conducted in PubMed and CENTRAL to identify studies including LT selection criteria of HCC patients.

**Study Selection:** The selected studies had LT selection criteria based solely on pre-LT variables. They included a minority of down-staged patients, over 80% of deceased donors, presented recurrence-free survival curves (or equivalent) with the number of patients at risk at different times, and had a follow-up period of over 3 years.

**Data Extraction and Synthesis:** Data was extracted from recurrence-free survival curves using a validated algorithm and subsequently used to calculate accuracy measures. The Preferred Reporting Items for Systematic Reviews and Meta-Analyses (PRISMA) reporting guidelines were applied.

**Main Outcome(s) and Measures(s):** Sensitivity, specificity, positive predictive value (PPV), negative predictive value (NPV), and accuracy at 3- and 5-years post-LT.

**Results:** Of 815 records screened, only 17 met our study inclusion parameters, embodying 14 LT selection criteria. All LT criteria achieved an adjusted PPV (aPPV) over 80%, indicating the correct selection of at least 80% of low-risk HCC patients. However, the adjusted NPV (aNPV) was below 50% in most cases, indicating that these criteria cannot correctly identify patients with a true high risk of recurrence. This raises major ethical concerns regarding the models’ ability to exclude patients from LT. Since a perfect model is nonexistent, we created a ranking to account for the distinct concerns of all stakeholders in LT eligibility in the context of HCC.

**Conclusions and Relevance:** These results highlight the urgent need for new tools/models with improved NPV to select more patients amenable to LT who are currently excluded. Whether through refined existing criteria or newly developed criteria, emphasis should be placed on specificity and NPV as key performance indicators in the emerging era of transplant oncology.

**Key points:**

- **Question:** What are the best criteria to select patients with hepatocellular carcinoma for liver transplantation?
- **Findings:** An objective ranking of existing criteria using accuracy measures considers the concerns of different stakeholders such as patients, physicians, payers, and organ allocation organisms. None of the analyzed criteria are ideal in satisfying all stakeholders in the selection of patients with hepatocellular carcinoma for liver transplantation. Criteria are even worse in accurately identifying patients who will not benefit from transplantation.
- **Meaning:** There is a need to refine current criteria by focusing on specificity and negative predictive value as key performance indicators.

## Introduction

Hepatocellular carcinoma (HCC) accounts for 75-85% of primary liver cancer and was the sixth most diagnosed cancer and the third leading cause of cancer-related deaths worldwide in 2022.^1^ Liver transplantation (LT) is considered the best curative treatment for HCC.^2,3^ However, its effectiveness is limited by organ availability, patient dropout during the waiting period, and post-transplant HCC recurrence.

Over the past decades, over 20 criteria have been developed to identify HCC patients who are less likely to experience HCC recurrence and would benefit most from LT^4–25^, ensuring fairer organ allocation. The most used models to assess recurrence risk after transplantation include the Milan, University of California San Francisco (UCSF), alpha-fetoprotein (AFP) score^26^, and Metroticket 2.0 (MT2.0) criteria^27^. These models are primarily based on tumour morphology^4,13,14,16^ with the AFP score and MT2.0 also considering tumour biology (AFP level).^4,14^ These criteria identify HCC patients with acceptable post-transplant 5-year recurrence risks as low as 15%^23,28–34^ and are associated with a 5-year survival rate of up to 70%. They effectively recognize patients with a low risk of recurrence (good prognosis). However, there is no consensus on which patient selection method is best. This lack of consensus is mainly due to: 1) limited number of prospective validations in external cohorts; 2) limited comparisons between different criteria within the same patient cohorts; 3) inclusion of confounding variables in studies (e.g., not distinguishing between living and deceased donors); 4) usage of outcome measures such as overall survival (OS) and recurrence-free survival (RFS) to compare between criteria, despite these measures being highly dependent on the patient cohorts, leading to potentially misleading results; and finally, 5) failure to consider the concerns of the main stakeholders (patients, physicians, payers, and organ-allocation organizations (OAOs)) in HCC-related LT decisions. Additionally, while all methods effectively select good prognosis patients (high PPV), their performance in identifying patients at high risk of relapse (bad prognosis) and the extent to which these tools wrongly disqualify patients with low risk of relapse for LT is unclear. In other terms, it is uncertain to what extent patients excluded from a transplant by these models would have experienced recurrence if they had received a transplant.

The lack of definitive criteria for good prognosis identification and their unclear performance in bad prognosis identification raises ethical questions. Therefore, there is a pressing need to compare existing criteria objectively using accuracy measures. These measures include sensitivity, specificity, positive predictive value (PPV), negative predictive value (NPV), and accuracy.^35^ These parameters are vital for prioritizing the strategies of various stakeholders involved in LT for HCC, ensuring that the accuracy ratings are directly associated with meaningful results.

The goal of this meta-analysis is to objectively evaluate different criteria for the selection of HCC patients for LT using accuracy measures, characterize each measure’s relevance for all LT stakeholders, and provide a baseline and framework for future studies.

## Methods

This study adhered to the Preferred Reporting Items for Systematic Reviews and Meta-Analyses (PRISMA) reporting guidelines.

### Literature Search

We conducted a systematic search in PubMed and CENTRAL (Cochrane Central Register of Controlled Trials) using the keywords: (“Liver” OR “Hepatic”) AND (“Transplant” OR “Transplantation”) AND (“HCC” OR “Hepatocellular Carcinoma”) AND “Selection Criteria” NOT “living donor” NOT “Downstaging” NOT “Resection”. We included studies defining HCC patients’ LT selection criteria up to June 2023. Additional eligible studies were screened from citations of relevant papers, mainly reviews and meta-analyses.

### Terminology/Definitions

RFS was defined as the time from LT to the first recurrence. Data from patients without documented recurrence at the last follow-up or death was censored. When terminologies/definitions were not explicitly stated in the publications, we assumed they were consistent with ours if separate plots for OS and RFS were presented (**eTable 1**).

### Inclusion Criteria

We used the PICOTS strategy to select literature studies, as outlined in **eTable 2**.

### Data Extraction

Data was extracted from recurrence/disease/tumour-free survival curves as reported elsewhere^36^. Briefly, a web plot digitizer (https://apps.automeris.io/wpd/) was used to extract the coordinates of each curve. These coordinates, along with the number of patients at risk at different time points, were used as inputs for the algorithm described by Guyot, *et al*^36^. The algorithm output provided individual patient data including follow-up time and recurrence status. In cases where the plots represented the cumulative recurrence rate/incidence/risk, each coordinate was subtracted from 1 before applying the algorithm. For studies covering both training and validation cohorts, data was extracted from the validation cohort only.

### Accuracy Measures

Accuracy measures were calculated as outlined in **eFigure 1**. Since PPV, NPV, and accuracy are influenced by event prevalence, they were adjusted to 0.87, the average of no-recurrence prevalence reported in the literature.^23,28–34^ This adjustment allows for a direct comparison of accuracy measures across different cohorts and time points. When criteria had multiple associated datasets (publications), the accuracy measures were calculated independently, and their means and respective standard deviations were presented. A positive outcome was defined as a patient who did not relapse and was within criteria.

### Risk of Bias Analysis

Each included study was assessed using the Tool to Assess the Risk of Bias in Cohort Studies (RBCS)^37^ and the Quality Assessment of Prognostic Accuracy Studies Tool (QUAPAS)^38^. The adaptation of both tools for the present study is described in **eTable 3**.

## Results

### Literature search results

Initially, 827 records were screened by title and abstract. After removing duplicates (n=12), non-English language studies (n=41), retracted studies (n=1), and studies outside the intended scope (n=570), the full text of 203 articles was assessed. Among these, inaccessible studies (n=3) and reviews and meta-analyses (n=41) were excluded, while their relevant citations were screened for additional eligible studies (n=29). Finally, 188 full-text articles were screened for eligibility (**Figure1**). For each criterion, 67% (n=125) and 55% (n=103) presented OS and RFS Kaplan-Meyer (KM) curves, respectively. Only 4% (n=8) of the articles compared criteria performance through accuracy measures, of which only 1% (n=2) stated the follow-up time. However, out of the 103 articles presenting RFS curves for a criterion alone, only 45% (n=46) met our eligibility standards by discriminating the number of at-risk patients (**eTable1**). Of these 46 studies, only 17 met all inclusion criteria and were included in the meta-analysis. These 17 articles represented 2% of the initially screened records, 8% of the eligible full-text articles, and 37% of the full-text articles with RFS curves and the number of at-risk patients (**Figure1**).

**Figure 1.**
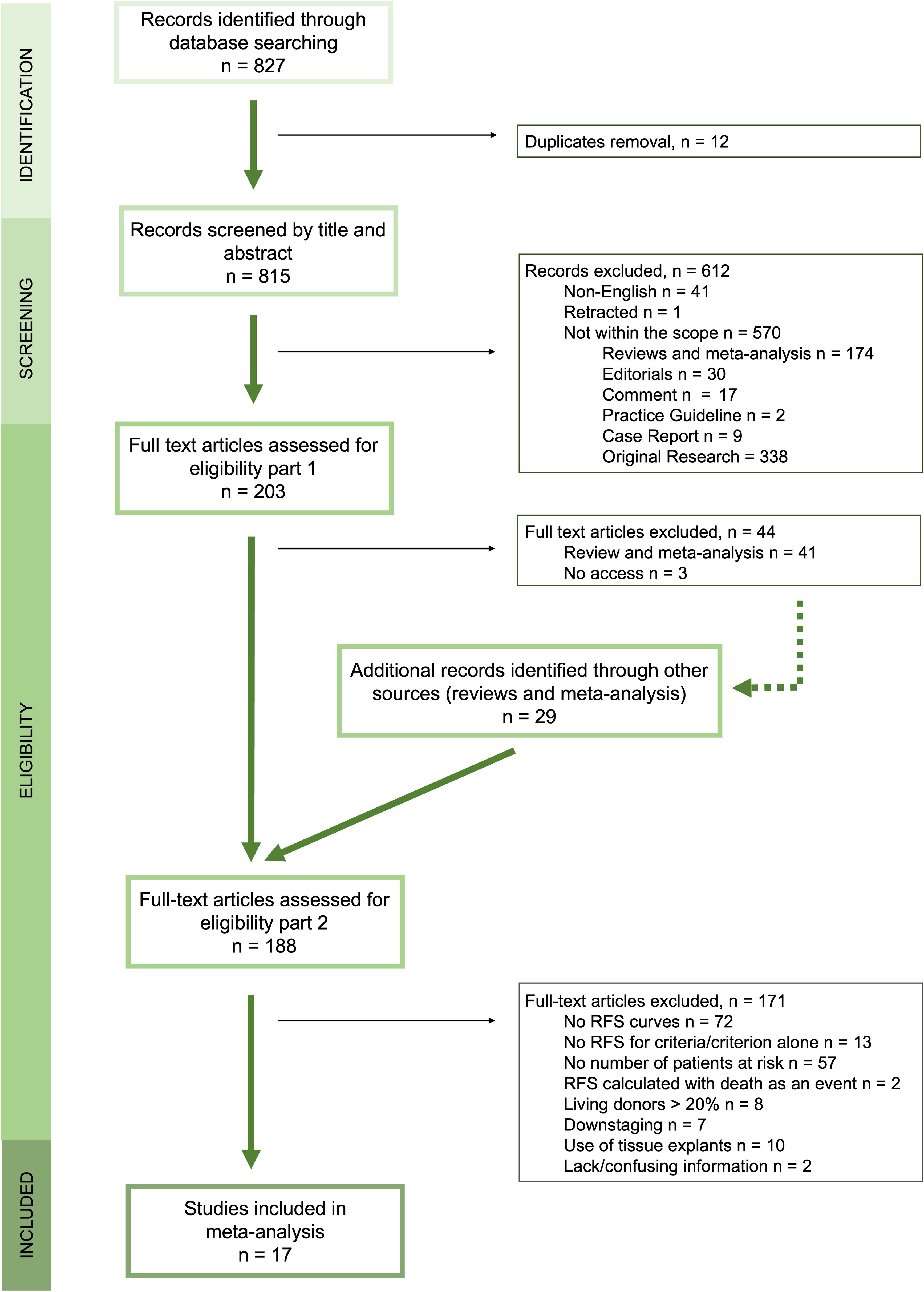
PRISMA flow diagram for study selection. Detailed information regarding each record is represented in **Supplementary Table 1.** RFS – Recurrence-free survival.

### Risk of Bias Assessment

Quality assessment of the 17 included studies used both RBCS and QUAPAS tools (**eTable3**). According to the results summary (**eTable4** and **eFigure2**), most studies presented a low risk of bias. However, for domains 7 and 12, 53% and 20% of the studies had a moderate and high risk of bias, respectively. This indicates that most studies (73%) did not follow patients for enough time. The censored cases were followed for less than 3 years in 50% of the studies and represented 30–60% of the cohort. Cohorts contained over 60% of censored cases in 20% of the studies. Additionally, concerning domain 8, 60% of the studies lacked information about pre-LT and post-LT therapies in the different groups (within and outside criteria), challenging the assessment of the real bias.

### Cohorts

This meta-analysis reviewed 17 articles to identify criteria for determining which patients would benefit from LT. Out of the 14 analysed criteria, the information for 11 (except Milan, AFP score and MT2.0) was obtained from one single study. **Table1** describes the selection criteria included in each model. A detailed description of the included studies is available in **eTable5**. Notably, well-known models like HALT-HCC (Hazard Associated with Liver Transplantation for Hepatocellular Carcinoma)^39^, pre-MORAL (Model of Recurrence After Liver Transplant)^6^, and NYCA (New York/California Score)^7,8^ were not included due to unmet inclusion criteria.

The LT criteria were at different stages of development, with some being tested in independent cohorts^17,22,23,25,40–46^, while others were in the validation (ArgScore and Warsaw criteria ^22,25,42^) or training (PLR, AFPdelta and GGT criteria ^18,19^) phase (**Table1**).

This study included data from 8032 patients, with the number varying between cohorts and decreasing with follow-up time due to an increasing number of censored cases (patients who died without recurrence or with lost follow-up). The recurrence rate also increased with the follow-up time (**eFigure3**).

### Accuracy measures

Accuracy measures were calculated using data from RFS curves at endpoints 3 and 5 years after LT without accounting for follow-up time and censored data. Within each criterion, our analysis found no significant differences in accuracy measures at different time points (**Figure 2**). Relapses occur mostly within the first three years (**eTable 5**), and the accuracy measures were normalized to a fixed value of no-recurrence prevalence described in the literature (0.87).^23,28–34^ **eFigure 4** displays the pre-normalization values. A clear trend for the NPV to be lower than all other accuracy measures was observed (**Figure 2** and **eFigure 4**).

**Figure 2.**
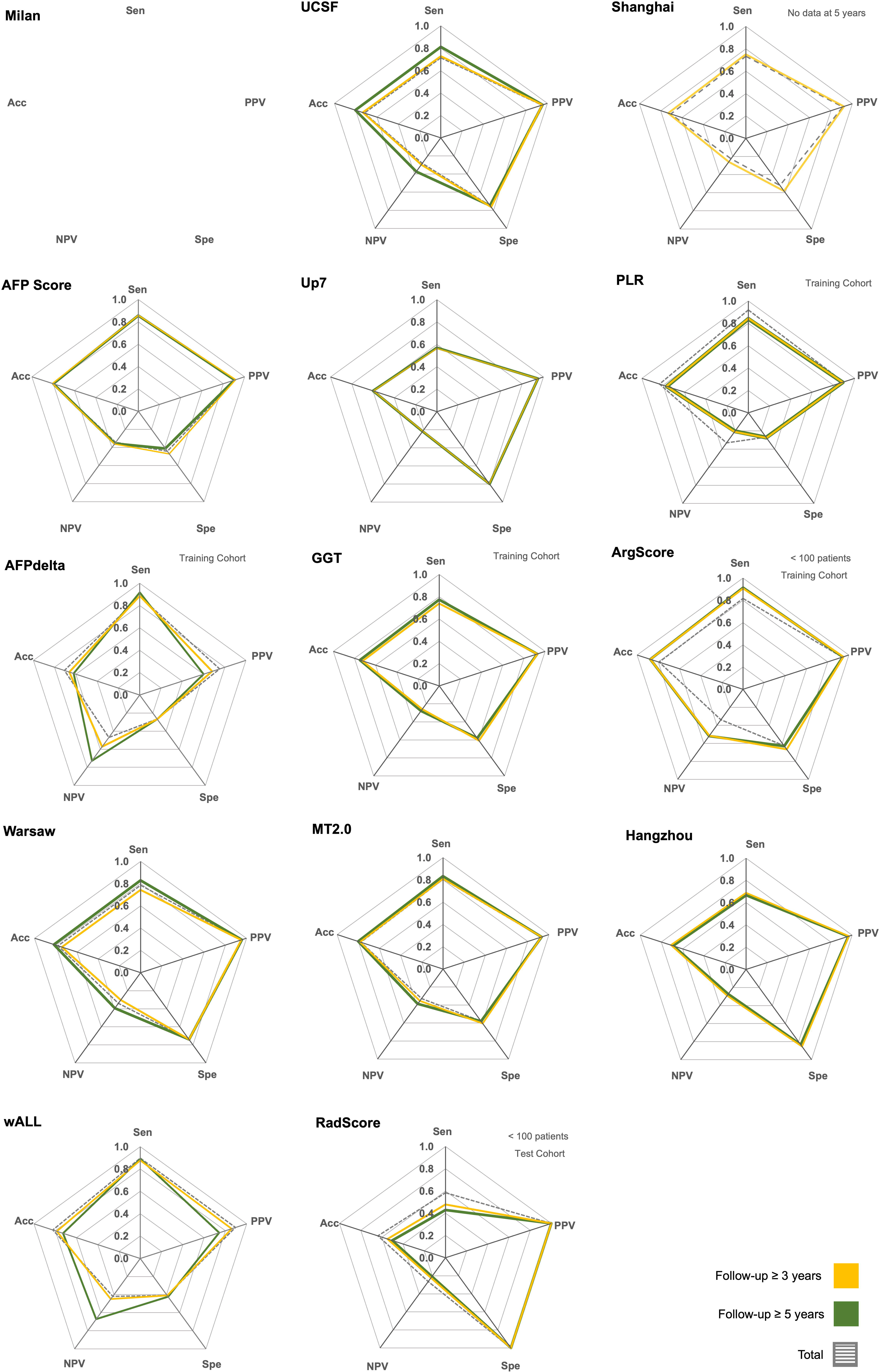
Accuracy measures for each criterion. Sensitivity (Sen) adjusted PPV (PPV), specificity (Spe), adjusted NPV (NPV), and adjusted accuracy (Acc) were calculated considering the mean no-recurrence prevalence described in the literature (0.87).

### The best criteria to include patients with a low risk of recurrence

Identifying patients with good prognosis, including those who likely will not relapse (true positives) and minimising their exclusion (false positives) is crucial. This requires high sensitivity/recall (ensuring that all no-relapse patients are within criteria) and a high PPV/precision (ensuring that all patients within criteria do not recur). When attributing the same weight to sensitivity and aPPV, only the ArgScore criteria showed both measures above 0.90 at both endpoints (**Figure 3A** and **3B**), making it the best option. The wALL, AFPdelta, AFP score, PLR, and MT2.0 criteria followed, presenting both measures above 0.80 at both endpoints.

**Figure 3.**
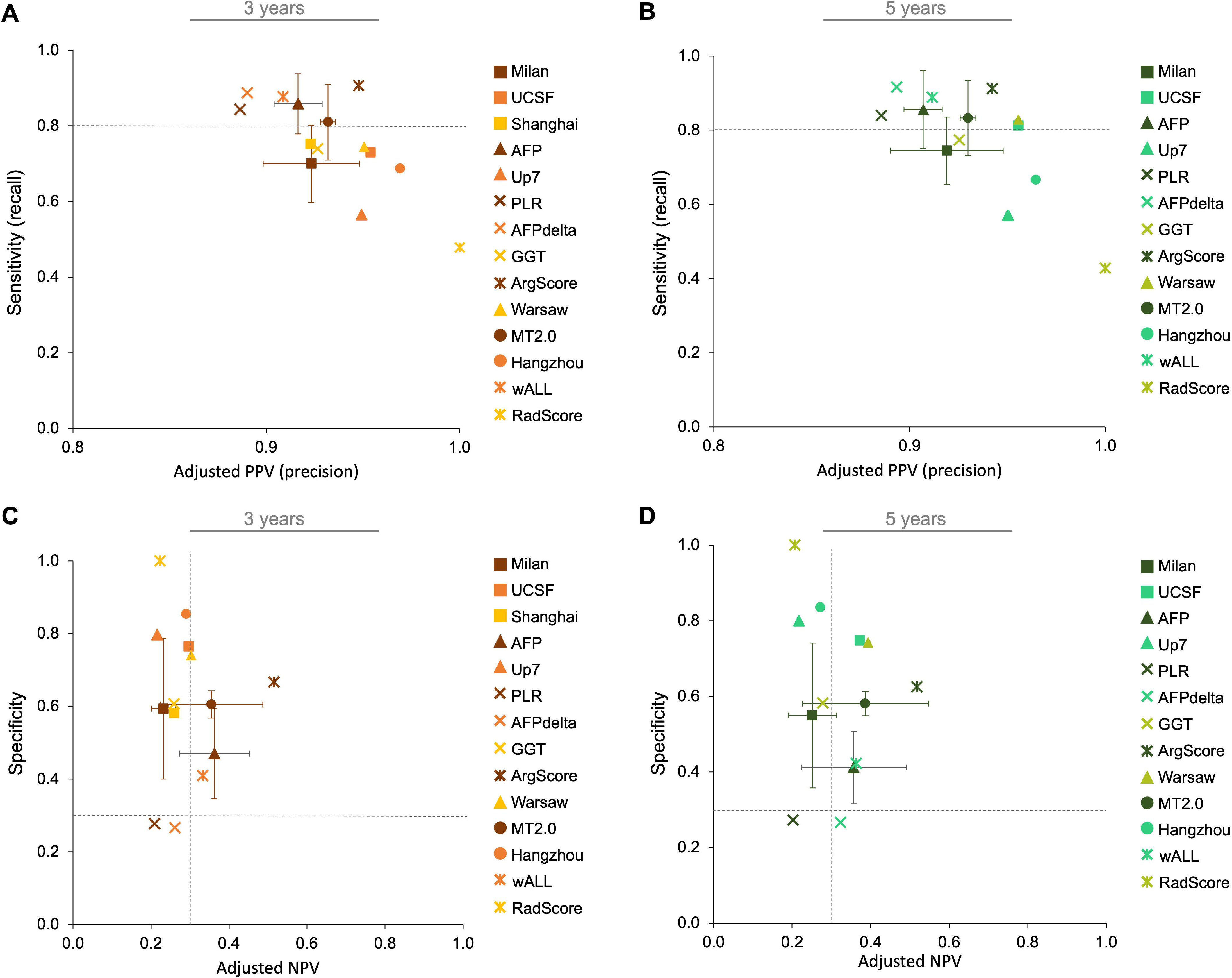
The Best criteria to include and exclude patients with good and bad prognosis after LT, respectively. The best criteria to include patients with good prognosis after LT should present a high sensitivity and adjusted PPV at 3 years **(A)** and 5 years **(B)** after LT. In contrast, the best criteria to exclude patients with bad prognosis after LT should present a high specificity and adjusted NPV at 3 years **(C)** and 5 years **(D)** after LT. Criteria identified with (X) represent cohorts with < 100 patients or training or validation cohorts. All accuracy measures were calculated considering the mean of no-recurrence prevalence described in the literature (0.87)

### The best criteria to exclude patients with high risk of recurrence

To correctly identify patients with poor prognosis it is necessary to maximize the exclusion of those at risk of relapse (true negatives) while minimizing the inclusion of those who will not (false positives). Criteria with high specificity (accurately pinpointing the patients who will relapse outside criteria) and high NPV (ensuring that most patients outside the criteria will relapse) are best for excluding patients who will relapse after LT. The ArgScore was the best criterion for correctly excluding patients from LT, with accuracy measures above 0.50 (**Figure 3C** and **3D**). It was followed by UCSF, AFP score, wALL, Warsaw, and MT2.0 criteria, all presenting a specificity and negative predictive value of at least 0.30, at 3 (**Figure 3C**) and 5 years (**Figure 3D**).

### Best criteria combining inclusion and exclusion of patients for LT

To define the best criteria for selecting patients for LT, the risk of relapse after the procedure should be minimized by including low-risk and excluding high-risk patients, a concept known as adjusted accuracy (aAcc). The ArgScore has the highest aAcc (0.88), followed by wALL, AFPdelta, and AFP score with aAcc of at least 0.80 at both endpoints (**Figure 2**).

### The best criteria for meeting the different stakeholders’ needs in LT

The main stakeholders in the context of HCC and LT are patients, physicians, payers, and OAOs. **Figure 4** illustrates the associations between accuracy measures and stakeholders’ priorities, along with the top six (top6) criteria for each parameter. The best criterion for each stakeholder was determined based on its frequency within the top6 ranking for each accuracy measure. Physicians prioritize increased accuracy in their decision-making process of selecting or denying an LT to a patient. For both patients and physicians, criteria with high NPV are desirable to avoid incorrectly denying a potentially life-saving transplant to a patient who would benefit from it. From the patientߣs perspective, criteria associated with high sensitivity are crucial to ensure that most low-risk patients are within the range of LT criteria and thus eligible for transplantation. Differently, payers such as hospitals and insurance systems may prioritize higher PPV because it guarantees the success of the LT procedure by ensuring that most patients who receive the transplant do not relapse. Finally, OAOs’ concern about the limited availability of organs requires specificity maximization. High specificity gains relevance in the context of patients and OAOs to ensure that high-risk patients are excluded from LT.

**Figure 4.**
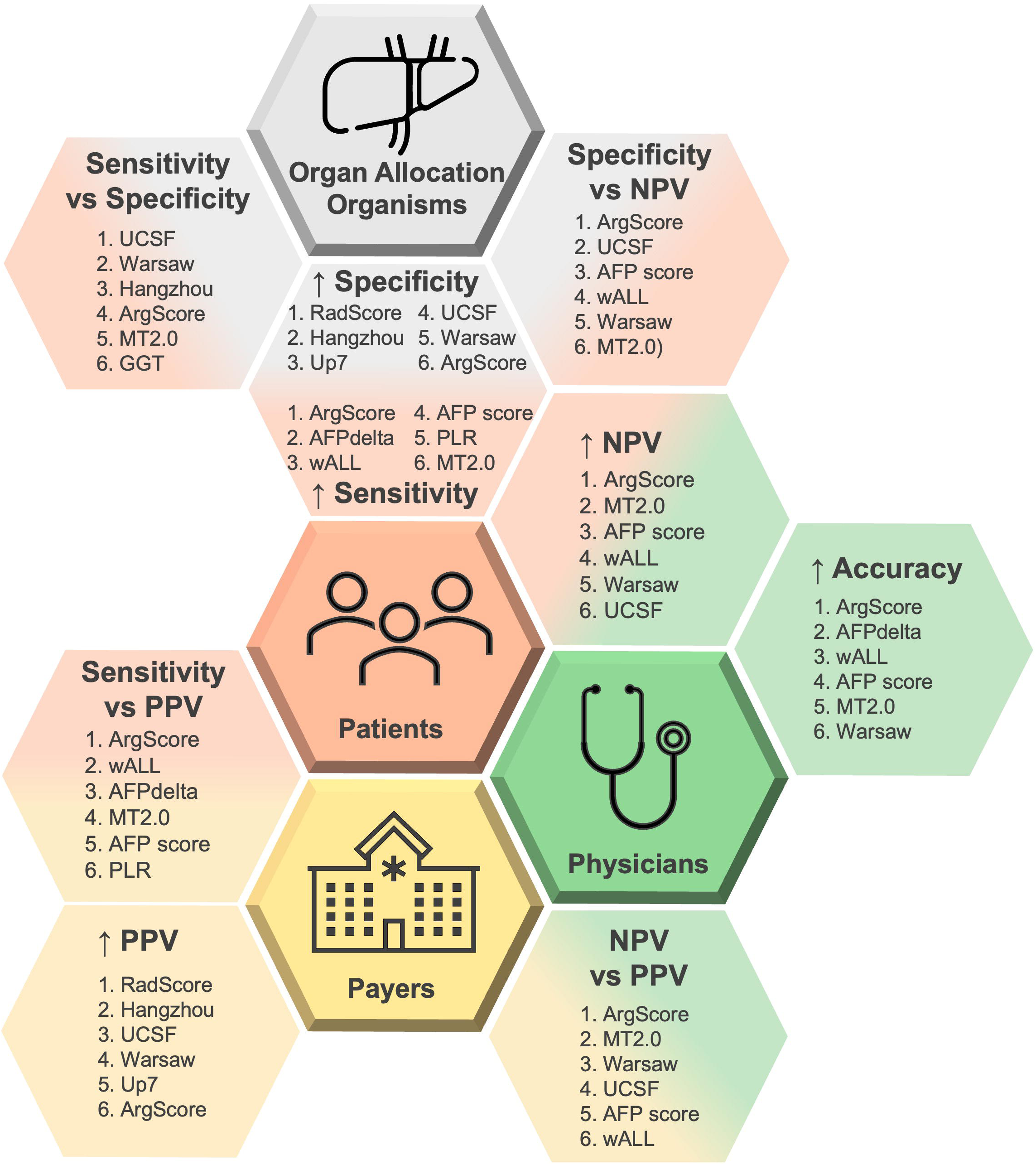
Accuracy measures in the clinical context. Representation of the four stakeholders in the clinical context of LT in HCC (patients, physicians, payers, and Organ Allocation Organisms) and their relationship with the different accuracy measures. Top 6 Criteria at both time points (3 and 5 years after LT) for each analyzed parameter. Where: NPV – negative predictive value; PPV – positive predictive value; UCSF – University of California, San Francisco; ArgScore – argentinian score; MT2.0 – metroticket 2.0; GGT – gamma-glutamyltranspeptidade; RadScore – radiological score; Up7 – up to seven; wALL – within all; AFP – alpha-fetoprotein; AFPdelta – AFP delta slope; PLR – platelet to lymphocyte ratio.

**Table 1.**
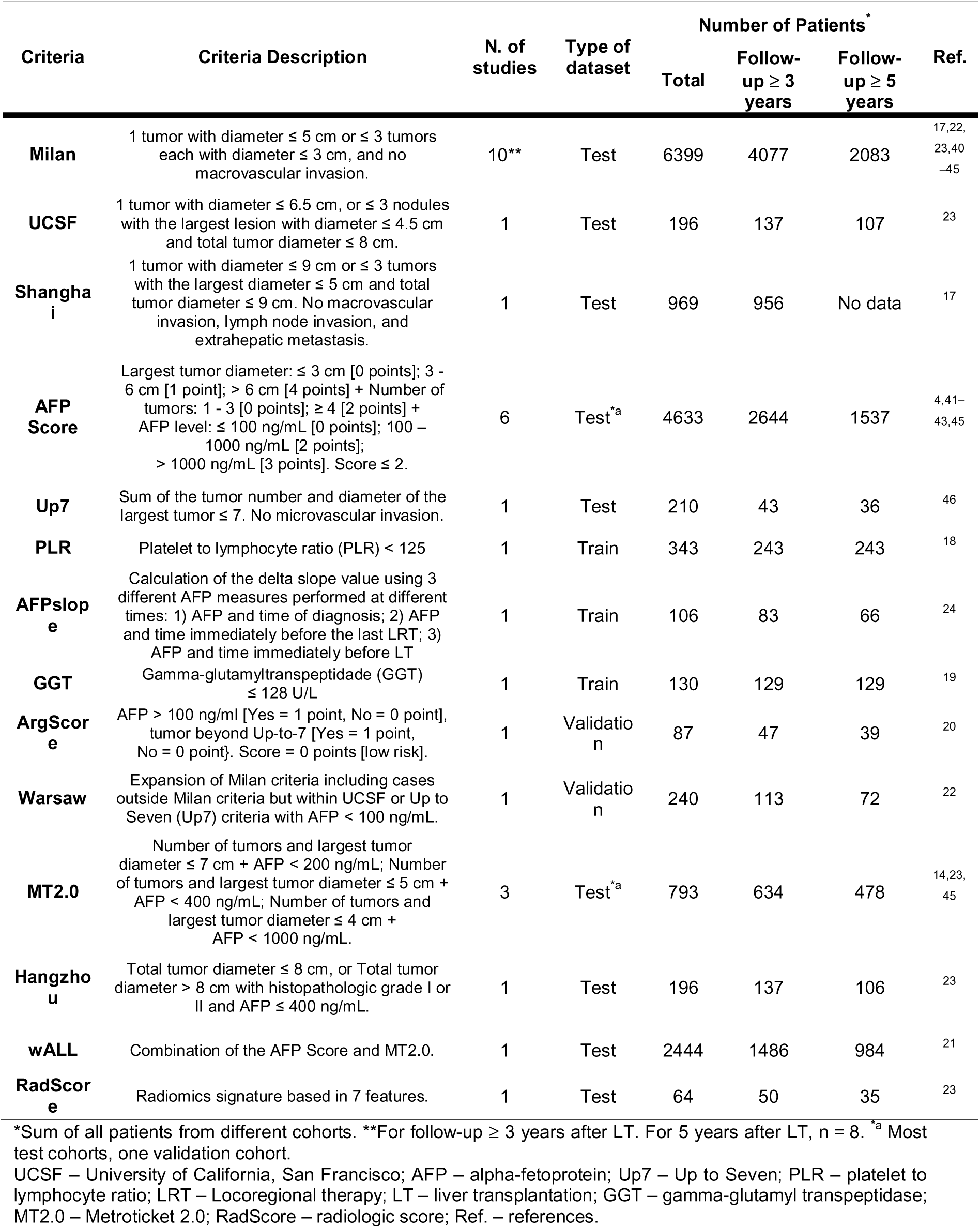
Data used in the meta-analysis structured by selection criteria.

In the scenario involving patients and OAOs, it is vital to simultaneously select criteria that are best at correctly identifying patients that will and will not relapse. This can be done by selecting the best-balanced interplay between specificity and sensitivity (**eFigure 5A** and **5B**). To address the concerns of both patients and physicians in identifying criteria that are best in denying an LT to a high-risk patient, the interplay between specificity and NPV must be analysed (**Figures 5C** and **5D**).

When analyzing each stakeholder individually, both ArgScore and Warsaw criteria were consistently present in all top criteria. Overall, when equally considering the concerns of all stakeholders as shown in **Figure 4**, the top6 criteria that consistently appear across the important measures for all stakeholders are ArgScore, MT2.0, Warsaw, UCSF, wALL, and AFP score.

## Discussion

This meta-analysis is the first to use accuracy measures to compare the prognostic value of various criteria for selecting HCC patients for LT with deceased donors. The results indicate that no perfect criteria exist, and the feasibility of each criterion depends on the stakeholders involved. Due to ethical concerns, such as avoiding the denial of the best standard of care to patients, international guidelines favour criteria that focus on sensitivity and/or PPV, promoting the inclusion of patients for LT, rather than excluding patients who would not benefit from LT. This preference is reflected in the results, with NPV being the accuracy measure with the lowest value. The low NPV associated with all analysed criteria means that a substantial number of HCC patients are still wrongly excluded from LT, which is unacceptable from both the patient’s and the physician’s perspectives. This situation calls for the urgent development of new predictive models with higher specificity and NPV. While we provide a ranking of the criteria based on their alignment with the diverse stakeholders’ needs, the ideal tool should equally balance all stakeholders’ concerns. Not all studies, some involving other criteria, met our inclusion standards due to inconsistent or incomplete reporting data, so results should be interpreted cautiously.

Despite the absence of a perfect criterion, the strict literature inclusion standards we used in this meta-analysis allowed for calculating accuracy measures for each criterion and comparing the performance results of many currently implemented criteria. In general, the results showed that when equally considering the concerns of all stakeholders, the top6 criteria were ArgScore, MT2.0, Warsaw, UCSF, wALL, and AFP score. Interestingly, Milan was the only criterion not included in the top6 among the most clinically adopted criteria^27^. Thus, the benchmark position of Milan criteria should be reconsidered in favour of more recent and properly validated models such as UCSF, wALL, MT2.0, or AFP score. While ArgScore and Warsaw criteria also presented good performance metrics, their results should be carefully interpreted since they lack validation in independent datasets (only one validation or one training).

We used accuracy measures and focused on RFS as the outcome event to directly compare different models. We chose RFS over OS as the outcome event because it is directly linked to poor patient survival^32–34^, places a higher cost on healthcare systems, and does not include causes unrelated to cancer. Interestingly, only 61.7% of the eligible papers included RFS KM curves, excluding reviews and meta-analyses. Among the 61.7%, less than half (37.9%) presented the number of at-risk patients in at least three different time points, allowing for data extraction. Of these studies, only 36.4% were included, as the others had confounding variables such as a living donor rate above 20%, downstaging cases above 50%, or using data from tissue explants. As a result, well-known criteria such as HALT-HCC^39^, pre-MORAL^6^, and NYCA^7,8^ were excluded from this meta-analysis. To enable a more comprehensive comparison of all developed criteria, future studies reporting on criteria performance should include RFS KM curves, with clear discrimination of the number of patients at risk at different time points and/or accuracy measures for different time points and indicating what was considered an event.

New clinical contexts are now arising in several European countries (e.g., Spain, and Italy) where the number of available organs for LT is increasing due to the reduction of other LT indications such as hepatitis C. In this scenario, we should anticipate that more patients will be considered for LT with expanded criteria. According to this study’s results, such criteria should also focus on increasing NPV and specificity. We propose two main ways to achieve this: 1) validate existing models through retrospective studies in historical cohorts or prospective studies either in living donation or downstaging contexts; 2) incorporate tumour molecular characteristics, as a deeper understanding of HCC biology improves outcomes after LT^47^. Several studies exploring explant-based biology, have highlighted the potential prognostic value of distinct biological features such as allelic imbalance^48^, gene^5^ and micro-RNA^9,49,50^ expression profiles, and evolutionary distance^10^. These molecular features may be further supported by integration with AI approaches (e.g., machine learning) allowing for the analysis of large amounts of data to create prognostic algorithms optimized for specific accuracy measures^51,52^. An example is the HepatoPredict tool, which outperforms Milan and several other criteria by combining morphologic and molecular tumoral features through a machine-learning algorithm^5^. There is an urgent need to independently validate several good-performing criteria and/or develop new ones focusing on the correct exclusion of patients to complement the existing ones.

### Conclusion

Selecting LT candidates with a low risk of HCC recurrence is challenging. Current selection criteria have limitations, often compromised by incomplete patient data, potentially skewing outcomes. Accuracy measures allow objective comparison of current selection criteria, supporting a more substantiated prioritization of all stakeholders’ goals. Enhancements in these selection processes are necessary, with specificity and NPV being critical performance indicators. These parameters are increasingly important in the evolving LT landscape for HCC, influenced by the decline of viral diseases and the use of more effective tumour down-staging therapies. As organ availability increases and expanded LT criteria are adopted, a broader pool of patients will be considered for LT. This shift underscores an urgent need to refine existing LT criteria or develop new models incorporating tumour biology prognostic elements. Tools like HepatoPredict could be instrumental in this evolution, offering insights into future directions.

## Data Availability

The data analyzed during the current study are available from the corresponding author upon reasonable request.

## Acknowledgement

Authors’ contributions

CD, JPL, and LPF conceived the study. LPF designed and performed the study; analyzed and interpreted data; and drafted the manuscript. CD, JC, JPL, and LPF edited and revised the manuscript. All authors read and approved the final manuscript.

