## Supplementary Materials for "Role of accuracy measures in selecting hepatocellular carcinoma patients for liver transplantation"

**Supplementary Material:**

**eTable 1:** Supplementary Table 1.xlsx – Description of all records analyzed for the present meta-analysis.

**eTable 2 –** PICOTS strategy used to select literature studies. Studies not meeting these criteria, duplicate publications, studies in languages other than English or including post-transplant variables were excluded.

| **Population** | Adult patients (≥ 17 years of age) with HCC undergoing LT with a minority of patients submitted to downstage, and with < 20 % of living donors. For papers with no information regarding the percentage of living donors, prevalence studies ^1–6^ were used to determine if the living donor LT rate was < 20 % in those populations. |
| --- | --- |
| **Intervention** | Liver transplantation. |
| **Comparator** | Criteria for the identification of HCC patients that will benefit from LT only using variables measured pre-LT (e.g.: radiologically). |
| **Outcome** | RFS or cumulative risk/incidence/rate curves with the number of patients at risk for at least three different time points. RFS was preferred to OS since it has been directly related to poor patient survival ^7–9^, it represents a higher cost for healthcare systems, and it does not comprehend non-cancer-related causes. |
| **Timing** | RFS at 3 and/or 5 years after LT. |
| **Setting** | Any study design analyzing pre-LT variables except study protocols, reviews, meta-analyses, letters, case reports, experimental (animal models), conference abstracts, comments/opinions, editorials, and practice guidelines. |

**eTable 3 –** Adaptation of the Cochrane tool to assess the risk of bias in Cohort Studies and the Quality Assessment of Prognostic Accuracy Studies Tool (QUAPAS) in the present meta-analysis.

| **Domain** | **Tool** | **Name** | **Description considering the present meta-analysis** |
| --- | --- | --- | --- |
| **D1** | To Assess Risk of Bias in Cohort Studies | Population Bias | Was the selection of patients within and outside criteria drawn for the same population? If yes, a low risk of bias was considered. |
| **D2** |  | Exposure bias | Confidence in the exposure (within or outside criteria) assessment. If the selection of patients as within or outside criteria were based on secure records (e.g.: CT scan or MRI), a low risk of bias was considered. |
| **D3** |  | Outcome of interest at start of study bias | If it is possible to be confident that the outcome of interest (recurrence and non-recurrence) was not present at start of the study, a low risk of bias was considered. |
| **D4** |  | Prognostic variables between groups bias | If the same variables were analyzed in the two groups (within and outside criteria), a low risk of bias was considered. |
| **D5** |  | Presence of prognostic factor bias | If it is possible to be confident in assessing the prognostic factors (e.g., number of tumors and diameter), a low risk of bias is considered. Confidence is associated with the use of robust databases, |
| **D6** |  | Assessment of outcome bias | If it is possible to be confident in the outcome (recurrence and non-recurrence) using, for example, medical records, a low-risk bias was considered. |
| **D7** |  | Follow-up bias | Assess if the follow-up of cohorts was adequate considering the amount of missing data. When a cohort showed < 30% of patients censored before 3 years of follow-up, it was classified as low-risk bias. When the rate of those patients varied between 30 – 60 % a moderate risk bias was considered. Finally, cohorts with > 60 % of patients censored before 3 years were classified as high risk of bias. Furthermore, when the percentage of censored patients before 3 years varied more than 50 % between groups (within and outside criteria), a level of bias was added. |
| **D8** |  | Co-intervention bias | Assess if the co-interventions (such as pre-LT and post-LT therapies) were similar between groups (within and outside criteria). For differences until 10 % - low risk of bias; between 10 – 30 % - moderate risk of bias; and over 30 % - high risk of bias. |
| **D9** | QUAPAS | Participant recruitment bias | Participant selection method. Similar to D1. |
| **D10** |  | Index test bias | If the index test (criteria) was applied to all participants in the same way, a low risk of bias was considered. |
| **D11** |  | Target event bias | Assessment of outcome. Similar to D6. |
| **D12** |  | Study flow bias | Assessment of patients lost during follow-up and time horizon from index test to outcome. Similar to D7. |
| **D13** |  | Analysis bias | If the statistical methods used were appropriated. When a data representation considering the occurrence of recurrence and the censored data was presented, a low risk bias was considered. |

**eTable 4 –** Assessment of risk of bias of the studies included in the meta-analysis

| **Study** | **Criteria** | **D1** | **D2** | **D3** | **D4** | **D5** | **D6** | **D7** | **D8** | **D9** | **D10** | **D11** | **D12** | **D13** |
| --- | --- | --- | --- | --- | --- | --- | --- | --- | --- | --- | --- | --- | --- | --- |
| Toso_08 ^10^ | Milan | Low | Low | Low | Low | Low | Low | Moderate | Low | Low | Low | Low | Moderate | Low |
| Fan_09^11^ | Milan | Low | Low | Low | Low | Low | Low | Low | No info | Low | Low | Low | Low | Low |
|  | Shanghai | Low | Low | Low | Low | Low | Low | Low | No info | Low | Low | Low | Low | Low |
| Duvoux_12^12^ | AFP score | Low | Low | Low | Low | Low | Low | Low | No info | Low | Low | Low | Low | Low |
| Lei_13 ^13^ | Up7 | Low | Low | Low | Low | Low | Low | Low | Low | Low | Low | Low | Low | Low |
| Lai_15 ^14^ | AFPdelta | Low | Low | Low | Low | Low | Low | Low | Low | Low | Low | Low | Low | Low |
| Xia_15 ^15^ | PLR | Low | Low | Low | Low | Low | Low | High | Low | Low | Low | Low | High | Low |
| Fu_16 ^16^ | GGT | Low | Low | Low | Low | Low | Low | Low | Low | Low | Low | Low | Low | Low |
| Notarpaolo_16 ^17^ | AFP score | Low | Low | Low | Low | Low | Low | Moderate | No info | Low | Low | Low | Moderate | Low |
|  | Milan | Low | Low | Low | Low | Low | Low | Moderate | No info | Low | Low | Low | Moderate | Low |
| Piñero_16 ^18^ | AFP score | Low | Low | Low | Low | Low | Low | High | Low | Low | Low | Low | High | Low |
|  | Milan | Low | Low | Low | Low | Low | Low | High | Low | Low | Low | Low | High | Low |
| PiñeroF_16 ^19^ | ArgScore | Low | Low | Low | Low | Low | Low | Moderate | No info | Low | Low | Low | Moderate | Low |
| Sapisochin_16 ^20^ | Milan | Low | Low | Low | Low | Low | Low | Low | High | Low | Low | Low | Low | Low |
| Grat_17 ^21^ | Milan | Low | Low | Low | Low | Low | Low | Moderate | Moderate | Low | Low | Low | Moderate | Low |
|  | Warsaw | Low | Low | Low | Low | Low | Low | High | Low | Low | Low | Low | High | Low |
| Grat_20 ^22^ | AFP score | Low | Low | Low | Low | Low | Low | Moderate | No info | Low | Low | Low | Moderate | Low |
|  | MT2.0 | Low | Low | Low | Low | Low | Low | Moderate | No info | Low | Low | Low | Moderate | Low |
|  | Milan | Low | Low | Low | Low | Low | Low | Moderate | No info | Low | Low | Low | Moderate | Low |
| Mazzaferro_18 ^23^ | MT2.0 | Low | Low | Low | Low | Low | Low | Low | No info | Low | Low | Low | Low | Low |
| Al-Ameri_19 ^24^ | AFP score | Low | Low | Low | Low | Low | Low | High | Low | Low | Low | Low | High | Low |
|  | Milan | Low | Low | Low | Low | Low | Low | High | Moderate | Low | Low | Low | High | Low |
| Piñero_23 ^25^ | AFP score | Low | Low | Low | Low | Low | Low | Moderate | No info | Low | Low | Low | Moderate | Low |
|  | Milan | Low | Low | Low | Low | Low | Low | Moderate | No info | Low | Low | Low | Moderate | Low |
|  | wALL | Low | Low | Low | Low | Low | Low | Moderate | No info | Low | Low | Low | Moderate | Low |
| Nie_23 ^26^ | Hangzhou | Low | Low | Low | Low | Low | Low | Moderate | No info | Low | Low | Low | Moderate | Low |
|  | MT2.0 | Low | Low | Low | Low | Low | Low | Moderate | No info | Low | Low | Low | Moderate | Low |
|  | Milan | Low | Low | Low | Low | Low | Low | Moderate | No info | Low | Low | Low | Moderate | Low |
|  | RadScore | Low | Low | Low | Low | Low | Low | Moderate | No info | Low | Low | Low | Moderate | Low |
|  | UCSF | Low | Low | Low | Low | Low | Low | Moderate | No info | Low | Low | Low | Moderate | Low |

Based on **eTable 3**. UCSF – University of California, San Francisco; AFP – alpha-fetoprotein; Up7 – Up to Seven; PLR – platelet to lymphocyte ratio; GGT – gamma-glutamyl transpeptidase;

MT2.0 – Metroticket 2.0; RadScore – radiologic score; Ref – references.

**eTable 5 –** Detailed description of the studies included in the meta-analysis.

| **Criteria** | **Reference** | **Number of patients** | | | **No-recurrence prevalence (%)** | | | **% Recurrence until 3 years after LT** | **Type of Dataset** |
| --- | --- | --- | --- | --- | --- | --- | --- | --- | --- |
|  |  | **Total** | **3 years after LT** | **5 years after LT** | **Total** | **3 years after LT** | **5 years after LT** |  |  |
| **Milan** | Toso_08 ^10^ | 275 | 112 | 69 | 89.8 | 77.7 | 59.4 | 89.3 | Test |
|  | Fan_09 ^11^ | 969 | 963 | N/A | 42.5 | 49.2 | N/A | 87.8 | Test |
|  | Notarpaolo_17 ^17^ | 573 | 372 | 261 | 86.0 | 82.5 | 71.3 | 81.3 | Test |
|  | Piñero_16 ^18^ | 327 | 182 | 118 | 84.7 | 76.4 | 57.6 | 86.0 | Test |
|  | Sapisochin_16 ^20^ | 504 | 426 | 351 | 81.0 | 81.2 | 74.6 | 83.3 | Test |
|  | Grat_17 ^21^ | 240 | 115 | 72 | 87.1 | 76.5 | 56.9 | 87.1 | Test |
|  | Grat_20 ^22^ | 282 | 184 | 122 | 82.6 | 79.9 | 59.8 | 75.5 | Test |
|  | Al-Ameri_19 ^24^ | 589 | 99 | N/A | 88.3 | 32.3 | N/A | 97.1 | Test |
|  | Nie_23 ^26^ | 196 | 138 | 106 | 53.6 | 35.5 | 14.2 | 97.8 | Test |
|  | Piñero_23 ^25^ | 2444 | 1486 | 984 | 84.7 | 80.3 | 65.1 | 74.2 | Test |
| **UCSF** | Nie_23 ^26^ | 196 | 137 | 107 | 53.6 | 35.0 | 15.0 | 97.8 | Test |
| **Shanghai** | Fan_09 ^11^ | 969 | 956 | N/A | 42.8 | 50.1 | N/A | 86.1 | Test |
| **AFP Score** | Duvoux_12 ^12^ | 435 | 326 | 75 | 89.4 | 88.3 | 40.0 | 82.6 | Validation |
|  | Notarpaolo_17 ^17^ | 574 | 366 | 253 | 86.2 | 82.8 | 70.8 | 79.7 | Test |
|  | Piñero_16 ^18^ | 325 | 181 | 121 | 84.9 | 76.2 | 59.5 | 87.8 | Test |
|  | Grat_20 ^22^ | 273 | 180 | 115 | 83.5 | 81.7 | 60.9 | 73.3 | Test |
|  | Al-Ameri_19 ^24^ | 589 | 100 | N/A | 88.3 | 32.0 | N/A | 98.6 | Test |
|  | Piñero_23 ^25^ | 2437 | 1491 | 973 | 84.7 | 79.9 | 64.6 | 73.3 | Test |
| **Up7** | Lei_13 ^13^ | 210 | 210 | 208 | 61.9 | 62.4 | 61.5 | 98.8 | Test |
| **PLR** | Xia_15 ^15^ | 343 | 243 | 243 | 57.7 | 42.0 | 41.2 | 97.2 | Training |
| **AFPdelta** | Lai_15 ^14^ | 106 | 83 | 66 | 71.7 | 63.9 | 54.5 | 80.2 | Training |
| **GGT** | Fu_16 ^16^ | 130 | 129 | 129 | 48.5 | 56.6 | 48.1 | 83.6 | Training |
| **NewScore** | PiñeroF_16 ^19^ | 87 | 47 | 39 | 81.6 | 68.1 | 59.0 | 93.8 | Validation |
| **Warsaw** | Grat_17 ^21^ | 240 | 113 | 72 | 87.1 | 76.1 | 56.9 | 87.1 | Validation |
| **MT2.0** | Grat_20 ^22^ | 276 | 181 | 117 | 83.0 | 80.7 | 59.8 | 74.5 | Test |
|  | Mazzaferro_18 ^23^ | 321 | 316 | 253 | 76.9 | 79.7 | 71.1 | 86.5 | Validation |
|  | Nie_23 ^26^ | 196 | 137 | 108 | 53.1 | 35.0 | 14.8 | 96.7 | Test |
| **Hangzhou** | Nie_23 ^26^ | 196 | 137 | 106 | 53.6 | 35.0 | 14.2 | 97.8 | Test |
| **wALL** | Piñero_23 ^25^ | 2444 | 1486 | 984 | 84.7 | 80.3 | 65.1 | 78.6 | Test |
| **RadScore** | Nie_23 ^26^ | 64 | 50 | 35 | 56.3 | 46.0 | 20.0 | 96.4 | Test |

UCSF – University of California, San Francisco; AFP – alpha-fetoprotein; Up7 – Up to Seven; PLR – platelet to lymphocyte ratio; GGT – gamma-glutamyl transpeptidase; MT2.0 – Metroticket 2.0; RadScore – radiologic score; Ref – references.

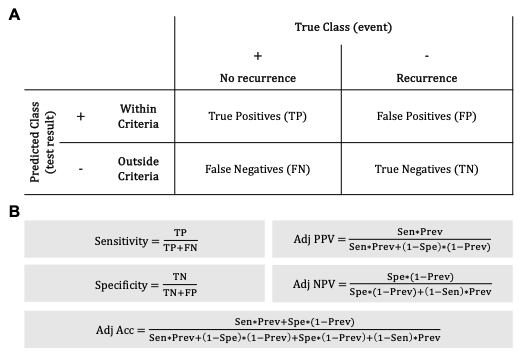

**eFigure 1 – Accuracy Measures Calculation.** The accuracy measures were calculated using the basis of a contingency table where the event was defined as no-recurrence and the positive test result was within criteria **(A).** Different formulas were used to calculate the accuracy measures. For the parameters dependent on event prevalence, the adjusted value was calculated **(B)**. In the present study, Sensitivity (Sen) concerns the number of patients who did not relapse and are within the criteria. Specificity (Spe) represents the number of patients who relapse and are outside criteria. Positive Predictive Value (PPV) and Negative Predictive Value (NPV) are the true positive and true negative results of a test, respectively and represent how likely it is if a specific criterion is used, to include or exclude a patient in case of no recurrence or recurrence, respectively^27^. Accuracy (Acc) represents the proportion of correctly classified patients – true positives (patients without recurrence within criteria) and true negatives (patients with recurrence outside criteria) – among all individuals. Since PPV and NPV concern the population to which the criteria are being applied^28^ and, together with Acc, are dependent on the no-recurrence prevalence^29,30^, a normalization was applied to all three parameters: Adjusted PPV (Adj PPV), Adjusted NPV (Adj NPV) and Adjusted Acc (Adj Acc).

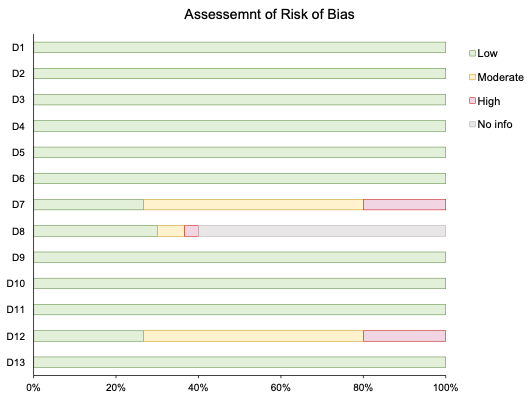

**eFigure 2 – Assessment of Risk of Bias of all studies included in the meta-analysis.** Each domain is described in eTable 3 and eTable 4.

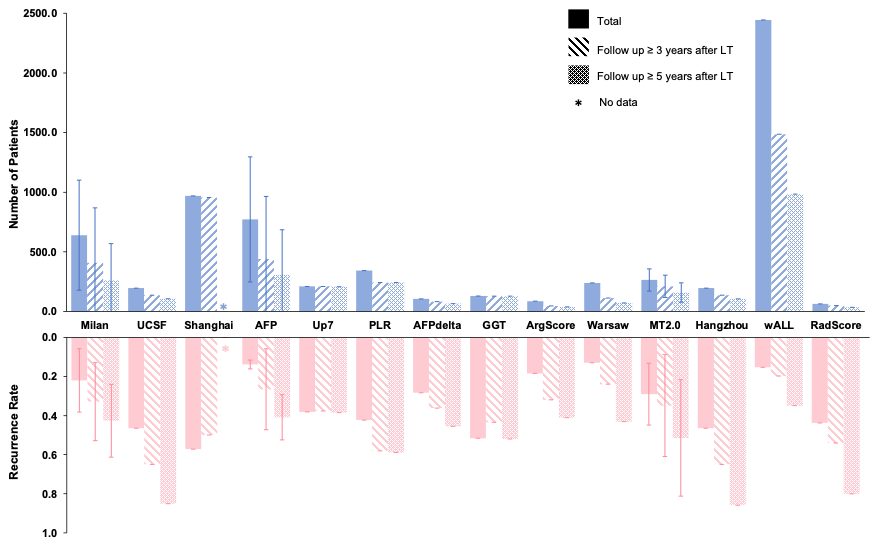

**eFigure 3 - Study Population.** Representation of the number of patients and respective recurrence rate for each criterion included in the study in total and at 3 and 5 years after LT.

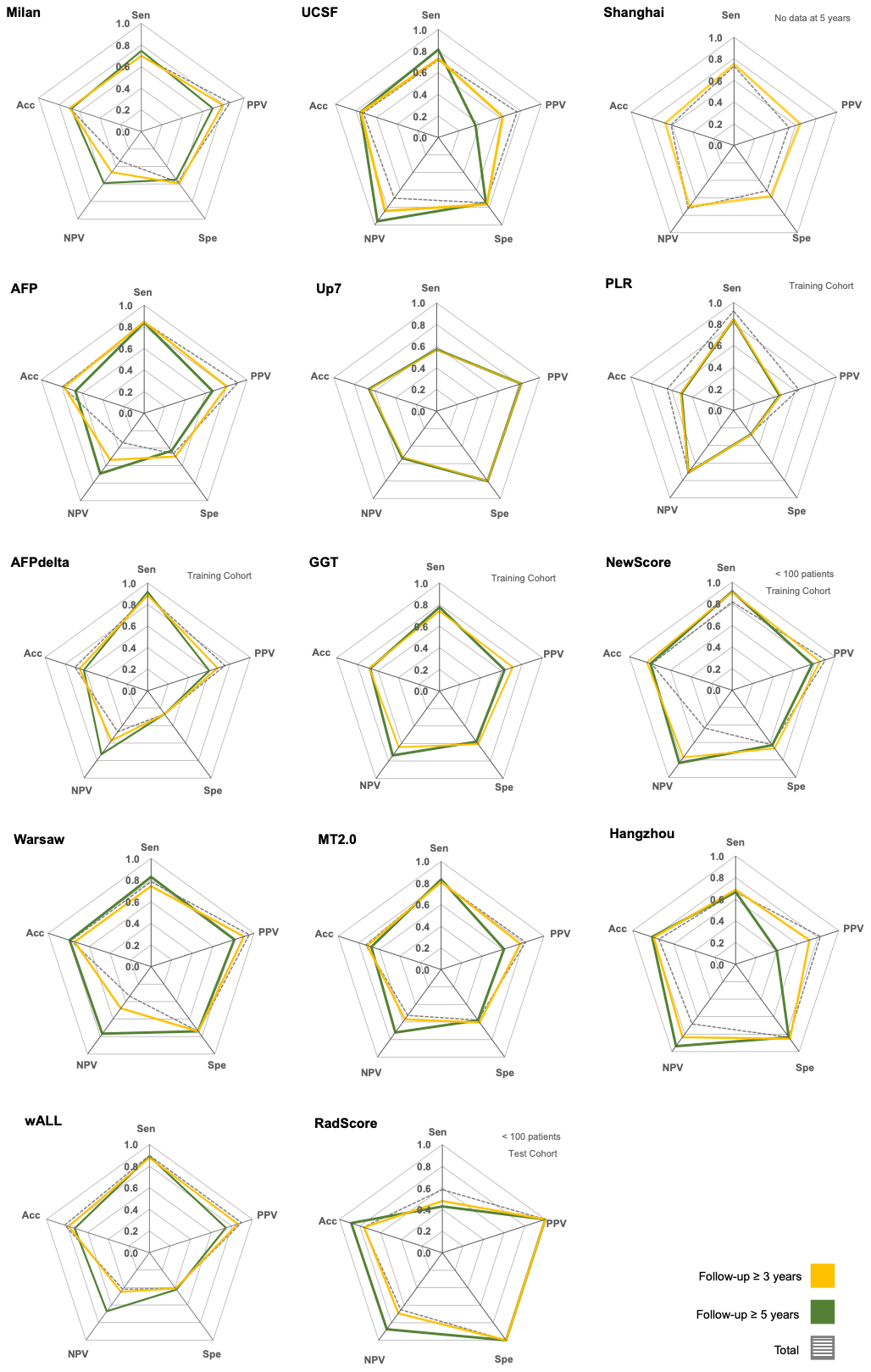

**eFigure 4 - Accuracy measures for each criterion without normalizations.** Sensitivity (Sen), positive predictive value (PPV), specificity (Spe), negative predictive value (NPV), and accuracy (Acc) were calculated directly from the independent datasets, i.e., not using a fixed prevalence value.

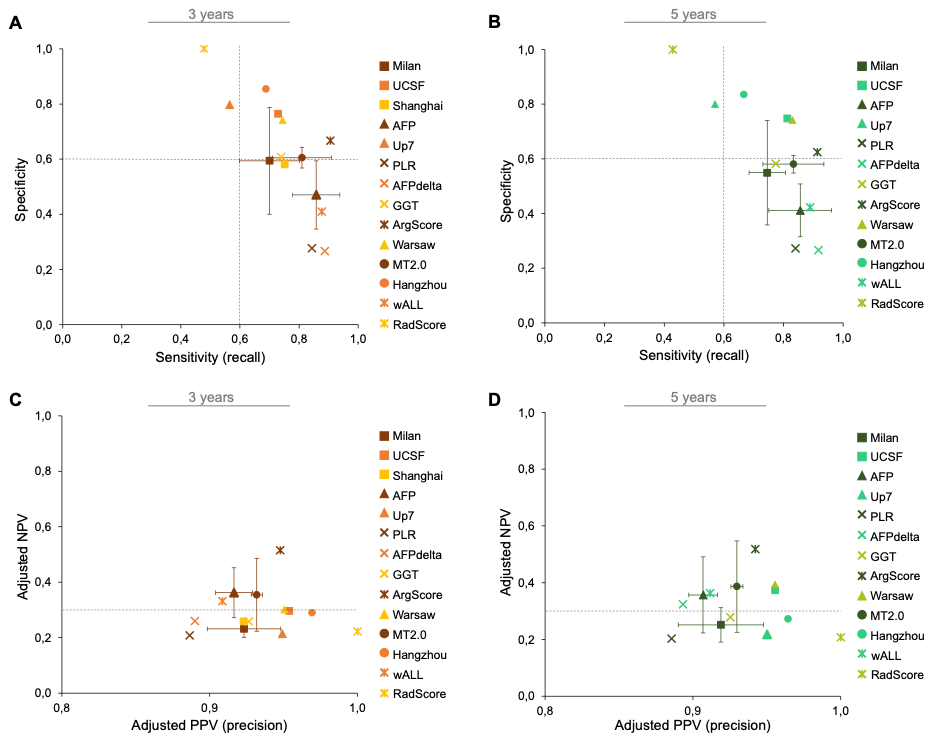
**eFigure 5 – The best criterion for patients’ classification.**

The best criterion to correctly classify patients that will and will not recur should present a high specificity and sensitivity at 3 years **(A)** and 5 years **(B)** after LT. In contrast, the best criterion to correctly include and exclude patients for LT should present a high adjusted NPV and adjusted PPV at 3 years **(C)** and 5 years **(D)** after LT. Criteria identified with (X) represent cohorts with < 100 patients or training or test cohorts. All accuracy measures were calculated considering the mean of no-recurrence prevalence described in the literature (0.87).
